# A model of COVID-19 transmission and control on university campuses

**DOI:** 10.1101/2020.06.23.20138677

**Authors:** Ben Lopman, Carol Y. Liu, Adrien Le Guillou, Andreas Handel, Timothy L. Lash, Alexander P. Isakov, Samuel M. Jenness

**Affiliations:** Emory University Rollins School of Public Health, Atlanta, GA 30322, USA; Department of Research and Public Health, Reims Teaching Hospitals, Robert Debré Hospital, Reims, France; College of Public Healh, University of Georgia, Athens, GA 30602, USA; Emory University School of Medicine, Atlanta, GA 30322, USA

## Abstract

In response to the COVID-19 pandemic, institutions of higher education in almost every nation closed in the first half of 2020. University administrators are now facing decisions about if and how to safely return students, staff and faculty to campus. To provide a framework to evaluate various strategies, we developed a susceptible-exposed-infectious-recovered (SEIR) type deterministic compartmental transmission model of SARS-CoV-2 among students, staff and faculty. Our goals were to support the immediate pandemic planning at our own university, and to provide a flexible modeling framework to inform the planning efforts at similar academic institutions. We parameterized the model for our institution, Emory University, a medium-size private university in Atlanta, Georgia. Control strategies of isolation and quarantine are initiated by screening (regardless of symptoms) or testing (of symptomatic individuals). We explore a range of screening and testing frequencies and perform a probabilistic sensitivity analysis of input parameters. We find that monthly and weekly screening can reduce cumulative incidence by 30% and 81% in students, respectively, while testing with a 2-, 4- and 7-day delay results in an 92%, 85% and 73% reduction in cumulative incidence in students over the semester, respectively. Smaller reductions are observed among staff and faculty. A testing strategy requires far fewer diagnostic assays to be implemented than a screening assay. Our intervention model is conservative in that we assume a fairly high reproductive number. We find that community-introduction of SARS-CoV-2 infection onto campus may be controlled with effective testing, isolation, contract tracing and quarantine, but that cases, hospitalization, and (in some scenarios) deaths may still occur. In addition to estimating health impacts, this model can help to predict the resource requirements in terms of diagnostic capacity and isolation/quarantine facilities associated with different strategies.

## Background

In an unprecedented response to the COVID-19 pandemic, schools (including institutions of higher education) in almost every nation closed in the first half of 2020.(1) For boarding institutions like universities, this involved both transitioning classes into online teaching as well as closing dormitories by sending students off-campus. School closure as a non-pharmaceutical intervention has been aimed at reducing contact among students, family members, teachers, and school staff.(2) It is thought to be an effective means of reducing disease transmission based on the understanding that younger people are important in transmission of respiratory viruses, like influenza. Closure of schools early in a pandemic is thought to be more impactful than delayed closing.(2) According to UNESCO, approximately 70% of the global student population has been affected, with closures of pre-school, primary, secondary, and higher education institutions.(1) Since SARS-CoV-2 infections are particularity severe among older adults while younger people still get infected and transmit,(3) university populations are unique in these degree of mixing across these age groups. Prior to the emergence of SARS-CoV-2, contact data on transmission of influenza, and other respiratory virus, provided the basis of current recommendations. Universities are important and unique in that they are frequently residential, involve students traveling long distances to attend, and are assets to their regional economies.

University administrators are now facing decisions regarding if and how to safely return students, staff and faculty to campus. As of the end of May 2020, approximately two-thirds of US universities are planning for in-person instruction for Fall 2020.(4) Universities considering campus re-opening need to estimate the resources necessary to interrupt and mitigate on-campus transmission by projecting the number of possible cases, needs for screening and testing, and boarding requirements for persons needing isolation and quarantine. To provide a framework to evaluate these questions, we developed a susceptible-exposed-infectious-recovered (SEIR) type of deterministic compartmental model. This model captures the transmission process and can therefore estimate the direct and indirect (transmission-mediated) effects of control strategies. For example, through model simulations, we estimated how testing and identifying SARS-CoV-2 infected students results in them being isolated, their contacts being quarantined, as well as all the infections averted by preventing the chains of transmission that would have otherwise occurred. Our goals were to support the immediate pandemic planning at our own university and to provide a flexible modeling framework to inform the efforts at similar academic institutions.

## Methods

We developed a dynamic model of transmission of SARS-CoV-2 among students, staff and faculty. We parameterized the model for our institution, Emory University, a medium-size private university in Atlanta, Georgia (numbers are for the main campus only). We expect the model could be applicable to other colleges and universities and therefore provide a public web-interface where key initial conditions and model parameters, such as student and staff population sizes can be varied (https://epimodel.shinyapps.io/covid-university/). The main features and assumptions are described in the following sections. Table 1 provides a full list of all parameter values, the model equations are shown in the appendix.

**Table 1.** Model parameters and ranges

| Parameter | Value | Range | Distribution | Symbol | Source |
| --- | --- | --- | --- | --- | --- |
| **Populations** |  |  |  |  |  |
| Total students | 15,000 |  |  |  | Univ. admin |
| Students living on campus | 4,500 | 1575 to 4500 | Uniform |  | Univ. admin |
| Staff and faculty | 15,266 |  |  |  | Univ. admin |
| **Natural history and clinical** |  |  |  |  |  |
| Latent period (days) | 3 | 2 to 4 | Gamma | $1/\alpha$ | (8) |
| Infectious period (days) | 7 | 6 to 8 | Gamma | $1/\gamma$ | (14) |
| Proportion severe - students | 0.0224 | 0.0133 to 0.0456 | Beta |  | (6) |
| Proportion severe - staff/faculty | 0.055 | 0.0327 to 0.1122 | Beta |  | (6) |
| Proportion fatal - students | 0.0006 | 0.0003 to 0.0014 | Uniform |  | (6) |
| Proportion fatal - staff/faculty | 0.0052 | 0.0029 to 0.0105 | Uniform |  | (6) |
| Proportion symptomatic - students | 0.35 | 0.27 to 0.43 | Beta | $1 - p_{asy,stu}$ | (15) |
| Proportion symptomatic - staff/faculty | 0.51 | 0.41 to 0.59 | Beta | $1 - p_{asy,saf}$ | (15) |
| **Transmission** |  |  |  |  |  |
| $R_0$: students to students | 2 | 0.7 to 2.5 | Uniform | $Ro_{stu,stu}$ | |
| $R_0$: on campus students to other on campus students | 1 | 0.3 to 1.4 | Uniform | $Ro_{on,on}$ | |
| $R_0$: Staff/faculty to student; staff/faculty to staff/faculty | 0.5 | 0.15 to 0.7 | Uniform | $Ro_{saf}$ | |
| Daily new cases in community (proportion) | 0.00005 | 0.000025 to 0.0001 | Beta | daily_new_case |  |
| Under-reporting factor for community infections | 10 |  |  | under_report |  |
| Efficacy of face-coverings and social distancing | 0.7 | 0.36 to 0.86 | Beta | eff_npi |  |
| **Testing and quarantine** |  |  |  |  |  |
| Time from onset of infectiousness to testing (1/days) | 2 | 7 to 1 | Uniform |  |  |
| Screening frequency (1/days) | 30 | 120 to 1 | Uniform |  |  |
| Duration of quarantine (days) | 14 | | | $1/\delta$ | |
| Number of contacts per case | 14 | 12 to 16 | Uniform | contacts | (16) |
| Proportion of contacts reached | 0.75 | 0.5 to 1 | Uniform |  |  |
| Proportion experiencing ILI symptoms per day | 0.00333 | 0.003 to 0.003667 |  |  | (10,11) |
| PCR sensitivity -- day 2 of infectiousness | 0.75 | 0.6 to 0.83 | Beta | $sen_2$ | (12) |
| PCR sensitivity -- day 4 of infectiousness | 0.8 | 0.7 to 0.85 | Beta | $sen_4$ | (12) |
| PCR sensitivity -- day 7 of infectiousness | 0.75 | 0.65 to 0.8 | Beta | $sen_7$ | (12) |

**Table 2.**
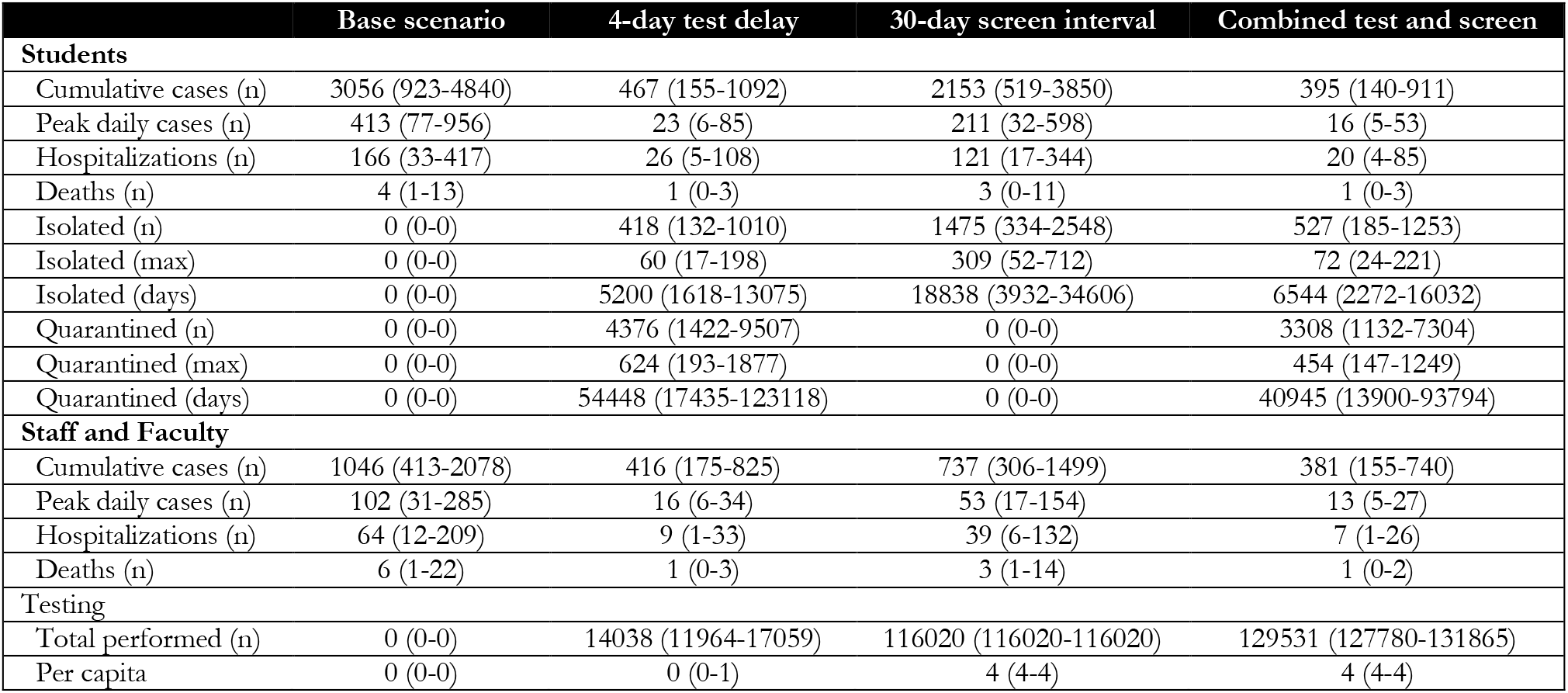
Cumulative outcomes at end of the semester on medium size university campus (approx. 15,000 students and 15,000 staff and faculty). Values are medians and 2.5 and 97.5^th^ percentiles of 1,000 model runs.

|  | Base scenario | 4-day test delay | 30-day screen interval | Combined test and screen |
| --- | --- | --- | --- | --- |
| **Students** |  |  |  |  |
| Cumulative cases (n) | 3056 (923-4840) | 467 (155-1092) | 2153 (519-3850) | 395 (140-911) |
| Peak daily cases (n) | 413 (77-956) | 23 (6-85) | 211 (32-598) | 16 (5-53) |
| Hospitalizations (n) | 166 (33-417) | 26 (5-108) | 121 (17-344) | 20 (4-85) |
| Deaths (n) | 4 (1-13) | 1 (0-3) | 3 (0-11) | 1 (0-3) |
| Isolated (n) | 0 (0-0) | 418 (132-1010) | 1475 (334-2548) | 527 (185-1253) |
| Isolated (max) | 0 (0-0) | 60 (17-198) | 309 (52-712) | 72 (24-221) |
| Isolated (days) | 0 (0-0) | 5200 (1618-13075) | 18838 (3932-34606) | 6544 (2272-16032) |
| Quarantined (n) | 0 (0-0) | 4376 (1422-9507) | 0 (0-0) | 3308 (1132-7304) |
| Quarantined (max) | 0 (0-0) | 624 (193-1877) | 0 (0-0) | 454 (147-1249) |
| Quarantined (days) | 0 (0-0) | 54448 (17435-123118) | 0 (0-0) | 40945 (13900-93794) |
| **Staff and Faculty** |  |  |  |  |
| Cumulative cases (n) | 1046 (413-2078) | 416 (175-825) | 737 (306-1499) | 381 (155-740) |
| Peak daily cases (n) | 102 (31-285) | 16 (6-34) | 53 (17-154) | 13 (5-27) |
| Hospitalizations (n) | 64 (12-209) | 9 (1-33) | 39 (6-132) | 7 (1-26) |
| Deaths (n) | 6 (1-22) | 1 (0-3) | 3 (1-14) | 1 (0-2) |
| **Testing** |  |  |  |  |
| Total performed (n) | 0 (0-0) | 14038 (11964-17059) | 116020 (116020-116020) | 129531 (127780-131865) |
| Per capita | 0 (0-0) | 0 (0-1) | 4 (4-4) | 4 (4-4) |

### Population Structure and Transmission

We modeled three distinct population groups with different degrees of interactions among them: students living on campus; students living off campus; and staff and faculty. We assume that staff/faculty can be infected by students and can infect other staff/faculty, with a reproduction number (before NPI) of 0.5. Student-to-student interactions leads to transmission at a higher rate, we assume a reproduction number (before NPI) of 2. We assume that students living on campus have a further increased transmission potential to other on-campus students, because congregate living is typical on most college campuses. We therefore assume those students infect on average 1 additional on-campus students.

Universities are planning an array of measures to limit transmission on campus. These may include mask-wearing; other personal protective equipment; smaller class sizes; staggered class times; enhanced cleaning protocols; enhanced hygiene; canceling large social gatherings; fewer students living in dorms and restricting off-campus movements. We lack data on the efficacy of all these interventions, especially in this specific population, but we assume that they will have an effect on transmission. We parameterize these non-pharmaceutical controls based on a systematic review of the effect of social distancing and face coverings (5)(and assume 50% compliance), and explore a range of values around this parameter.

Staff and faculty had a higher risk of severe illness and death (given infection) than students, based on accumulating evidence of age-differences in the case-fatality rate,(6) and then standardized using the student and staff/faculty age-structure at our institution. [For a full list of parameter values, see Table 1.] We further assume that a fraction of cases is asymptomatic and that, the probability of symptoms is greater for staff/faculty given their older age distribution than students. We assumed that asymptomatic infected persons are as infectious as those with symptoms; this assumption may overestimate the real transmission rate in this group.(7) We assume that infectiousness begins on the third day after infection; this latent period is shorter than the incubation period(8) to represent pre-symptomatic transmission.

We do not track transmission in the wider community, aside from incorporating a daily rate of introduction of virus onto campus from the community. In order to capture external infection from non-university sources, we modeled a constant daily rate of infection being introduced on campus. In our model parameterization, this is based on confirmed COVID-19 cases in Fulton and Dekalb Counties that surround our institution in early June (around 100 per day) and the combined population of the two counties.(9) We further assume that infection incidence is ten-times that of reported cases.(7) The model runs for a semester from the day classes start (August 26) to the end of term (December 19), or 116 days. We did not assume reduced transmission over traditional Fall or Thanksgiving breaks or consider alternative schedules.

### Intervention Design

In the model, control is initiated by SARS-CoV-2 diagnostics. Infected persons can be identified by reverse transcription polymerase chain reaction (RT-PCR) through either testing or screening, defined as follows. *Screening* is a strategy in which students, staff, and faculty are tested at a given frequency ranging from weekly to once per semester regardless of the presence of symptoms. *Testing* is a strategy whereby symptomatic students, staff, and faculty present for clinical care and are tested using RT-PCR. We assume a background level of persons with influenza-like symptoms caused by infections other than SARS-CoV-2, (10,11) who will test negative. Those with COVID-19 who test positive are immediately isolated. However, we assume that the diagnostic has imperfect sensitivity that varies based on what date of illness the test is performed.(12) There is evidence that PCR sensitivity varies over the course of infection, reaching a peak around day 7 of infection (or day 4 of infectiousness), then declines again. Therefore, we examined the impact of variation in the *testing* interval, defined as the average lag time between symptom onset and quarantine. Because infectiousness begins one day before symptom onset in the model, we simulated testing intervals ranging from a two -day to a one-week test delay. These testing scenarios are in the absence of any screening to isolate the causal effects of this more intensive intervention.

Following both screening and testing, those testing positive for COVID-19 were immediately isolated. Case isolation in the model mechanistically involved a complete reduction in their contact rate for the duration of infection. Positive test results also lead to contact tracing. Contact tracing is conducted by assuming public health authorities could elicit NA contacts per case detected with 75% of those successfully traced and quarantined. Quarantine, like isolation, was modeled as a complete reduction in the contact rate for the duration of infection. Some of those quarantined contacts might have been incubating but are now no longer able to infect since they are under quarantine.

### Parameterization and Analysis

In our base models, we simulated SARS-CoV-2 transmission and interventions for the Fall 2020 semester. Our main base model assumed NPI interventions but no screening or testing based-control. Counterfactual scenarios then varied the screening and testing rates, and the completeness of contact tracing. Our primary outcomes were both active cases per day and cumulative cases across the semester. The model tracked both total cases in each campus group (students versus staff/faculty) as well as severe cases and COVID-related mortality.

Given uncertainty in model parameters, we performed a probabilistic sensitivity analysis to determine the range of credible outcomes. In the probabilistic sensitivity analysis, we take 1,000 parameter draws using Latin Hypercube Sampling from the distributions in Table 1 and report the 2.5th and 97.5th centile of those runs (Appendix II). We use partial rank correlation coefficient to determine how much the modeled variation in cumulative incidence among students and faculty/staff depends on specific random parameters.

The model was built and simulated in the *EpiModel* package in the R statistical computing platform (13); the *LHS* package was used to perform Latin Hypercube Sampling. We also built an interactive web app for model exploration using the R Shiny framework. It can be accessed at https://epimodel.shinyapps.io/covid-university/.

## Results

We start by simulating transmission on campus (Figure 2) in which no diagnostic control measures are in place (no testing, isolation, contact tracing, or quarantine). With R0 of 3.5 for on-campus student and 2.5 for off-campus, case prevalence peaks at 807 cases (Range, 2.5th to 97.5th centiles: 263 to 1417) per day among students and 241 cases per day among staff/faculty (76 to 551), resulting in a cumulative of4150 (2449 to 5722) and 1744 (784 to 3176) cases at the end of the semester in a population of about 15,000 each. With our baseline levels of facemask and social distancing efficacy (70%) but with no diagnostics, we estimate case prevalence peaks at 413 (77-956) per day among students and 102 (31-285) cases per day among staff/faculty, resulting in a cumulative of 3056 (923-4840) student cases and 1046 (413-2078) staff cases at the end of the semester. Note that this number of symptomatic cases is substantially lower than the number of infections since we assume that 35% (27 to 43%) of infected students and 51% (41 to 59%) of infected staff are symptomatic, given infection.(15) We use this scenario as the baseline counterfactual for all subsequent comparisons.

**Figure 1.**
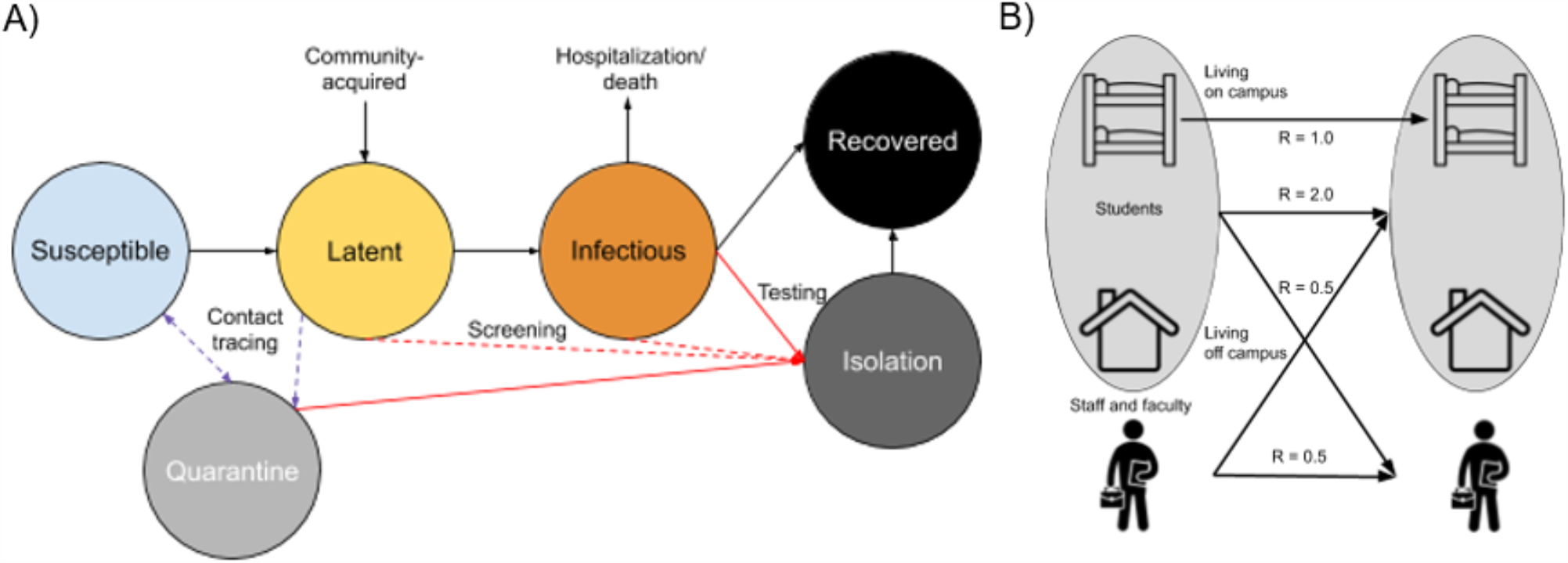
Schematic of A) Disease Structure and B) Student and Staff/Faculty Transmission Pathways

**Figure 2.**
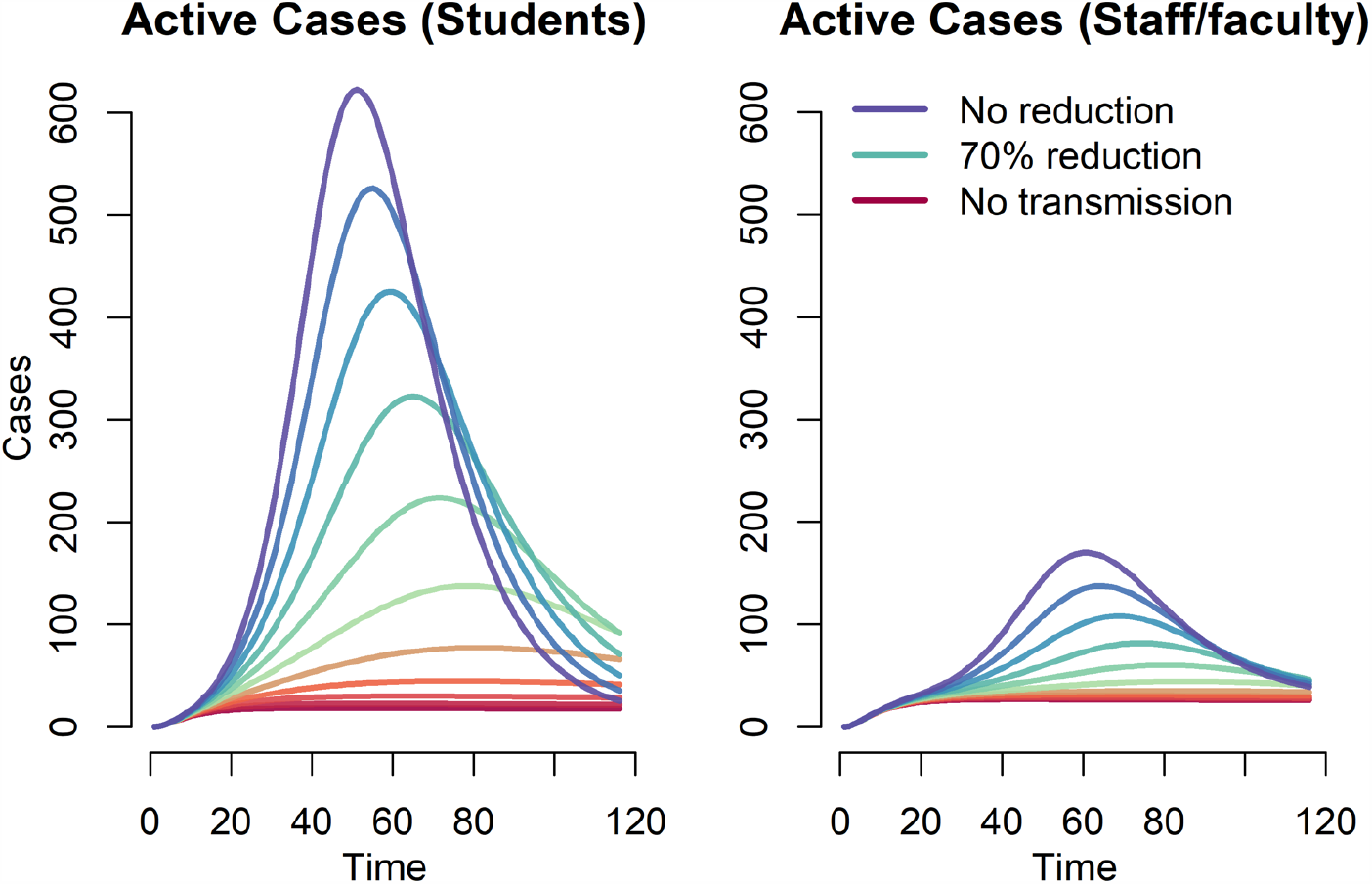
Effect of non-pharmaceutical interventions (with no testing and screening) on COVID-19 prevalence among students and faculty.

We next explored a wide range of screening intervals, from weekly to once during the semester (Figure 3). One-time screening, whereby the population is tested on average once during the 4-month semester, reduced cumulative student incidence overall by 8%; monthly and weekly screening reduced cumulative student incidence by 30% and 81% respectively. For staff and faculty, one-time screening reduced cumulative incidence by 9%; monthly and weekly screening reduce cumulative incidence by 29% and 65% respectively. For students, the cumulative incidence ranged from 583 (184-1595) with weekly screening to 2814 (809-4584) with one-time screening. For staff/faculty, the cumulative incidence ranged from 366 (144-751) with weekly screening to 950 (381-1895) with one-time screening.

**Figure 3.**
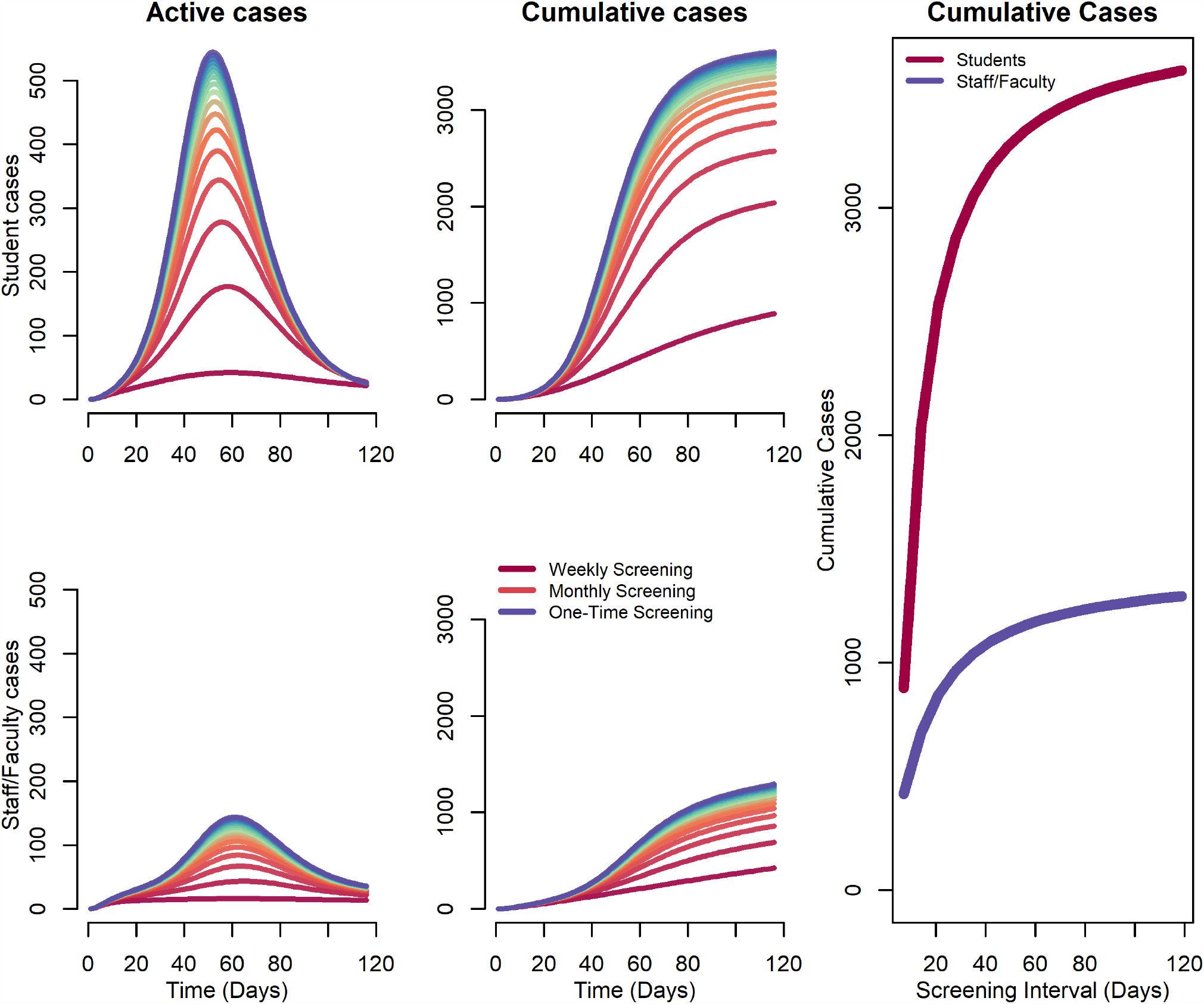
Impact of Screening Frequency on Projected Covid-19 Daily and Cumulative Incidence.

We then consider a testing-only strategy, which also includes contact tracing and quarantine. Assuming different delays between symptom-onset and receiving the tests, results are shown in Figure 4. We again plot the total number of cases among students and staff, with other parameter values at their base values. Here, with week-delayed testing (the least optimistic scenario), the expected cumulative incidence would be 826 (233-1792) for students and 503 (206-987) for staff/faculty. With a four-day delay testing interval, the expected cumulative incidence would be 467 (155-1092) for students and 503 (206-987) for staff/faculty. With a two-day delay testing interval, the expected cumulative incidence would be 250 (91-523) for students and 354 (144-700) for staff/faculty. These scenarios represent a 73%, 85% and 92% reduction in cumulative incidence over the semester among students and a 52%, 60% and 66% reduction in cumulative incidence among staff & faculty. Figure 3 also shows the general relationship between “contact tracing” success and cumulative incidence assuming either a 2-day, 4-day, or 7-day delay in testing/quarantine following symptoms. Although the testing interval can reduce the cumulative incidence, the greater impact of this testing scenario is achieved by the number of contacts reached.

**Figure 4.**
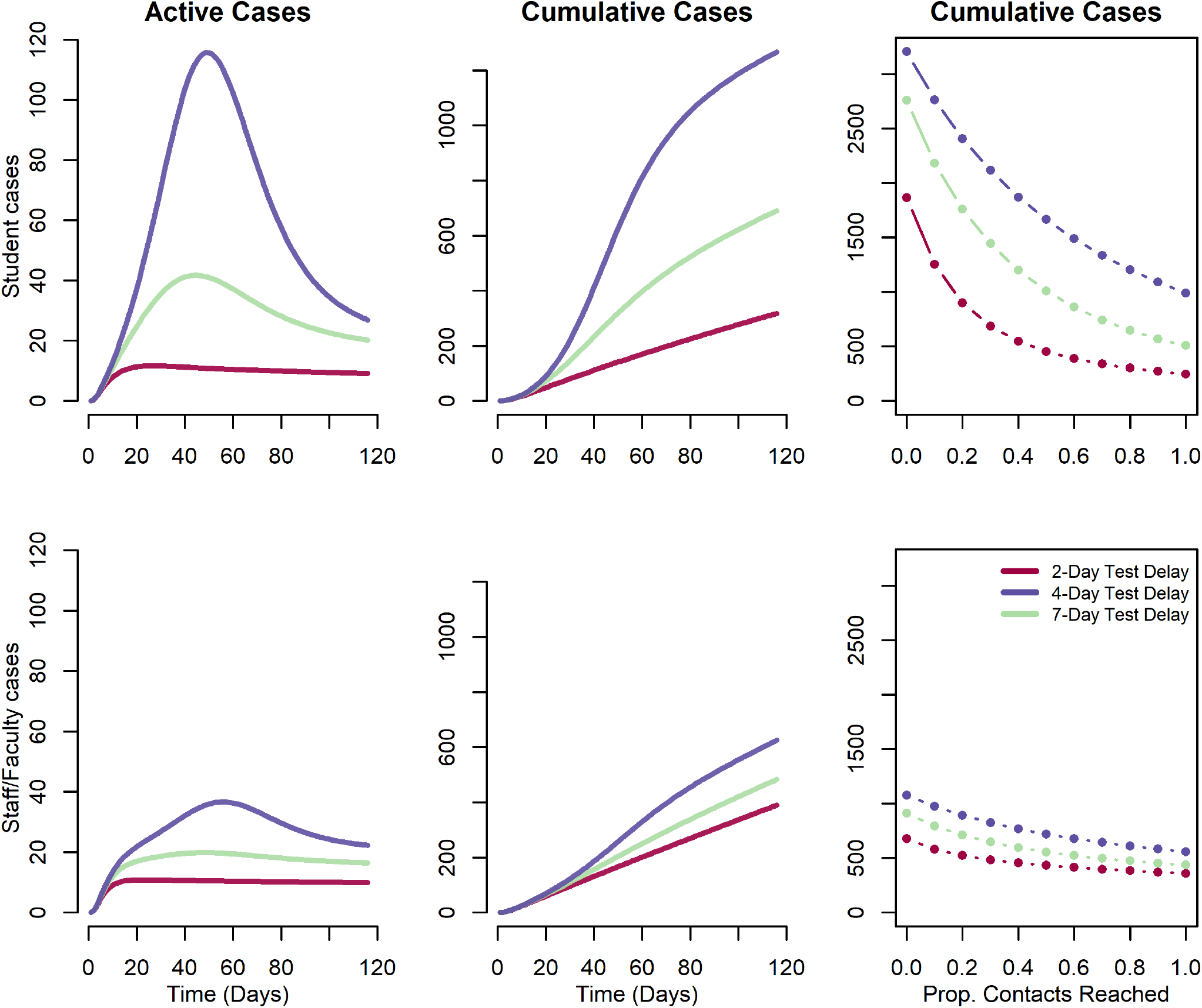
Impact of testing, contact tracing and quarantine at a range of testing delay intervals. Daily and cumulative Covid-19 incidence on university campus

In the final scenarios, we combined the testing and screening rates under different assumptions of contact tracing related to testing (Figure 5). Our model scenarios below varied the interval for screening between 1 week (7 days) and 1 semester (120 days) and testing at2-, 4- and 7-day delay, with the efficacy of contact tracing ranging from 0, 50% and 100%. These figure panels show cumulative incidence at the end of the semester for students only. When combined with testing, screening generally has little effect unless it is performed at least monthly.

**Figure 5.**
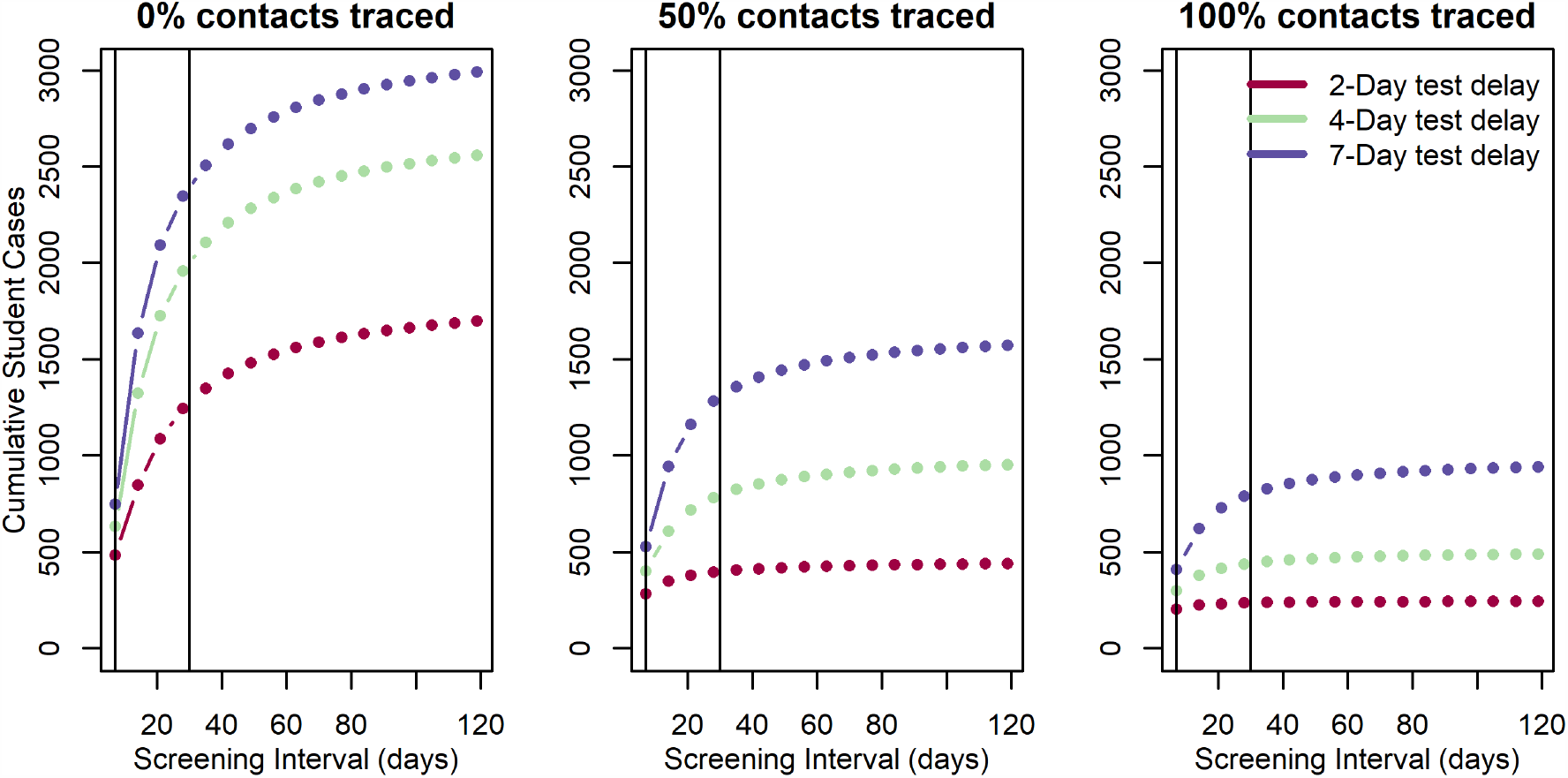
Impact of combined screening and testing of covid-19 cases among students. Vertical lines represent weekly and monthly screening.

**Figure 6:**
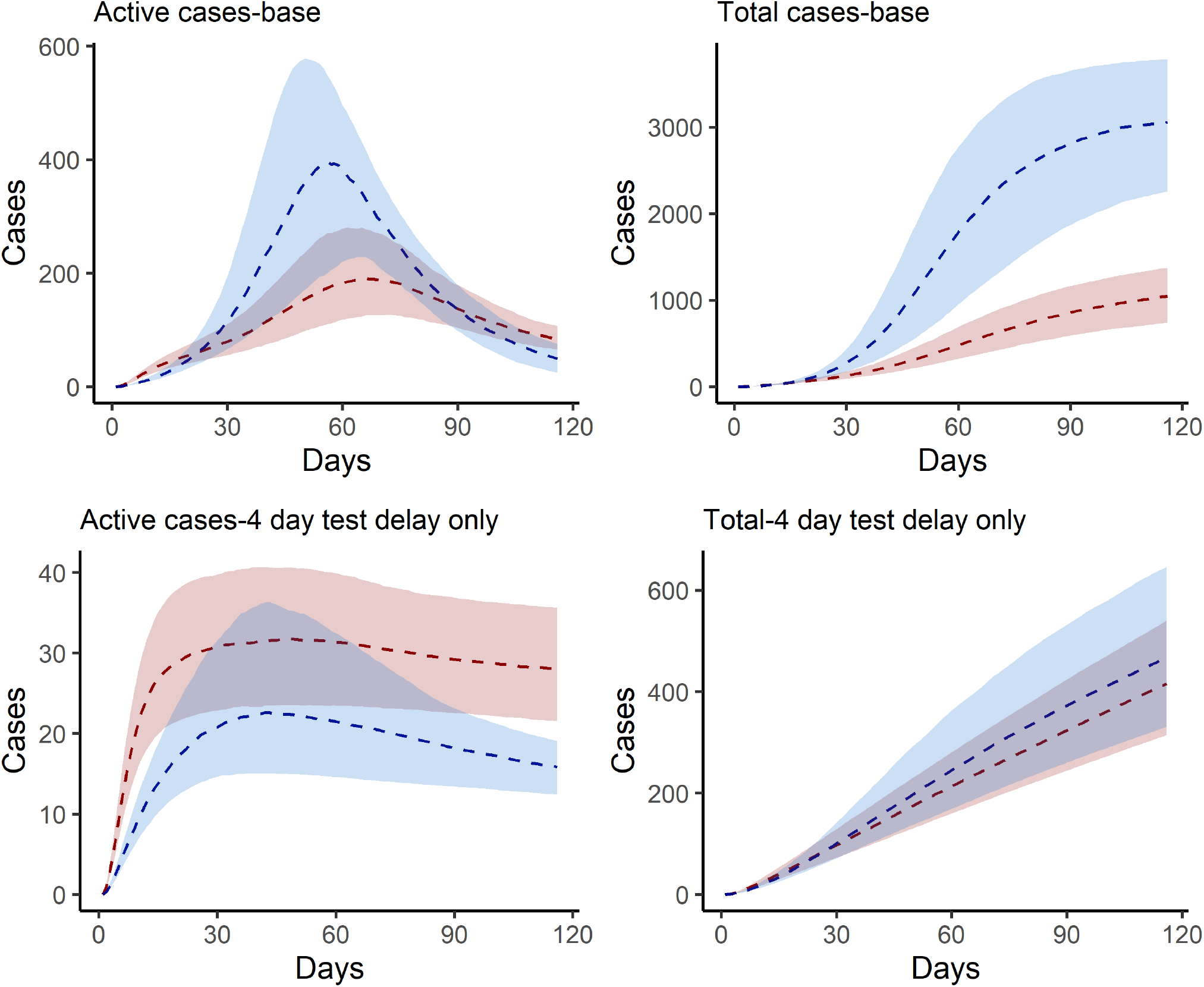
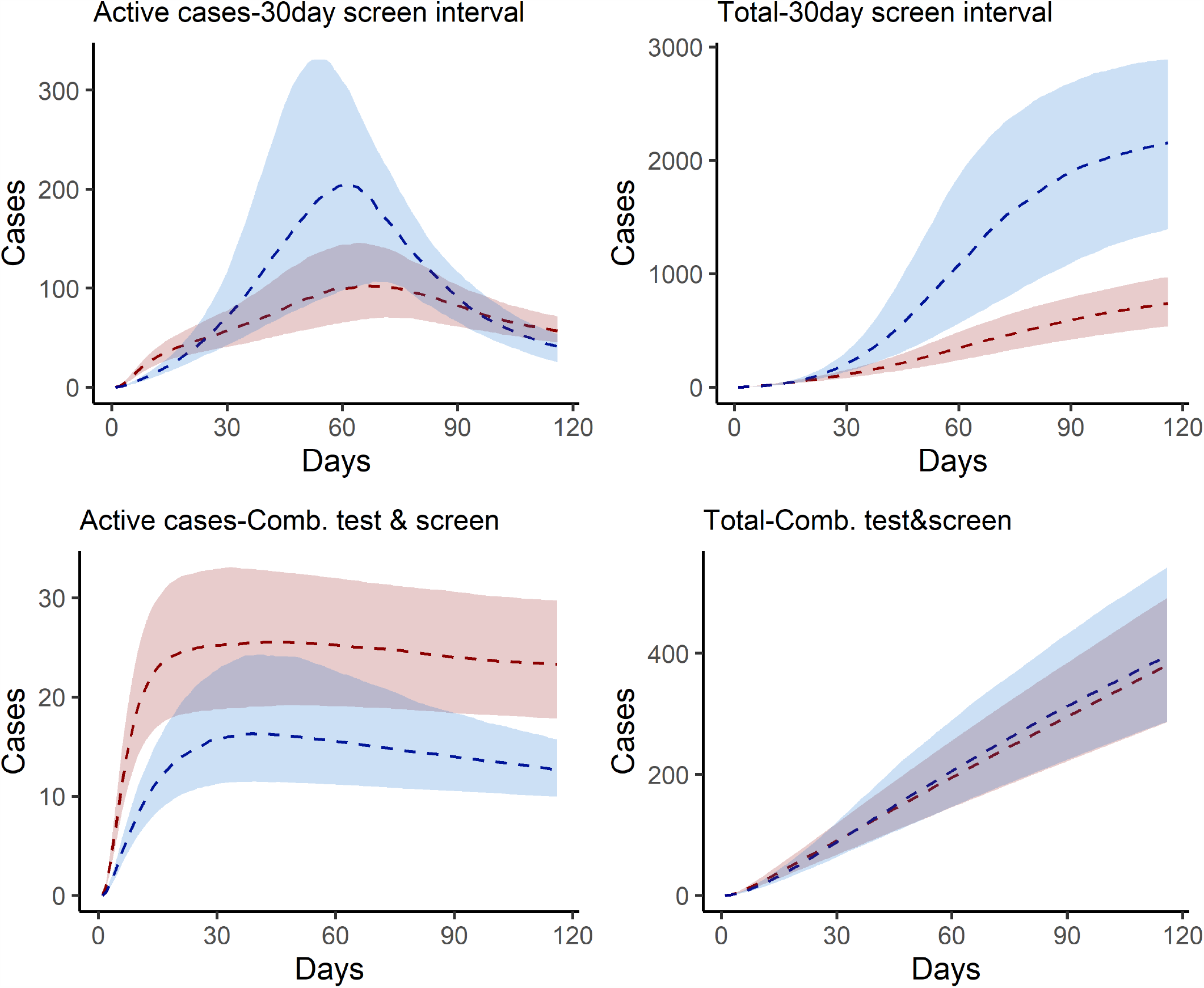
Estimated active and cumulative cases under intervention scenarios with 25th and 75th centile range.

**Figure 7:**
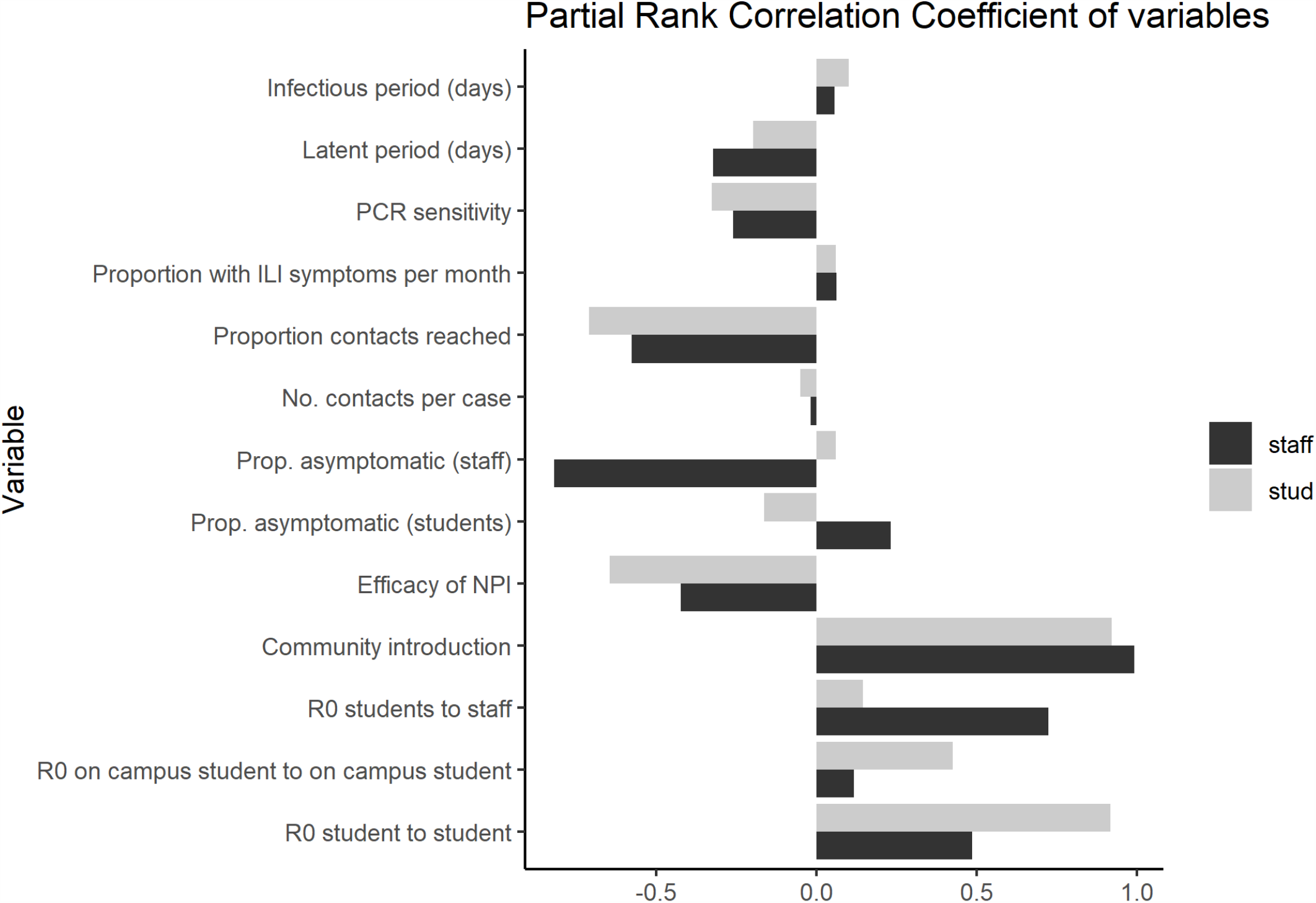
Partial rank correlation coefficient of key model inputs.

## Discussion

We find that unmitigated transmission of Covid-19 in a population of around 30,000 staff, faculty and students would lead to thousands of illnesses, many hospitalizations and likely deaths in this population, which is clearly an unacceptable outcome by administrators and the university community. Combined with measures to reduce transmission, a testing strategy whereby symptomatic students, staff and faculty are identified, administered viral testing, and isolated may be effective at controlling transmission. We find that the success of this strategy relies on contact tracing and quarantining most contacts of infected individuals. Screening would have to be performed at least monthly to have much of an impact on the course of the outbreak on campus and increases the sample collection and assay requirements considerably. However, because we assume the campus community is not a closed population and that there is an ongoing risk of importation of virus, there are considerable numbers of cases even under the most optimistic scenario, which therefore requires substantial resources. Overall, we recommend that these results be interpreted qualitatively, since there is considerable uncertainty in these projections stemming from lack of precision of parameter inputs (e.g. true R0 in this population).

There are a number of limitations to this modeling analysis, which we outline here. First, we lack empirical data about the efficacy of any prevention and control measures aside from testing that are implemented on campus. Smaller class sizes, staggered class times, use of face coverings, use of other protective equipment and general behavior change are not separately accommodated in this model.(5) If such data become available in campus population or ones that can serve as a good proxy, model parameters can be refined. Moving more students to off-campus housing has little effect on our projections because we make the assumption that transmission on-campus (R0 = 3.5) is only moderately higher than off campus (R0 = 2.5). This assumption is based on risk factor data on influenza-like illness among students during the 2009 H1N1 outbreak, but if more data become available, we could revisit this assumption.(17) In our model, the campus outbreak cannot go extinct because we assume a constant rate of introduction from the community. Depending on levels of student, staff and faculty behavior off-campus and the general prevalence in the surrounding community (Atlanta metro area in our model), this could be an under- or overestimate of risk. We have not explicitly included a scenario in which all or a subset of students (e.g., those residing on campus) are screened upon return to campus. Given our assumptions that student prevalence is the same as among the general population, screening on return would have limited effect, but would increase requirements by ∼4,500 to 15,000 tests, depending on the breadth of testing of the student body. Finally, we have not included seasonal effects whereby virus becomes more transmissible in Fall or alternative semester dates (e.g., end of classes at Thanksgiving break) whereby the period of campus transmission is reduced.

While we present numerical results for our univerity at a specific point in time, the model can be re-parameterized for other institutions and can be updated as the epidemiolgical situation shifts. We updated the model a number of times in discussions with our university leadership. Results from this framework have been influential in their ongoing decision-making. Community risk is a parameter that we updated as the local area’s incidence increased and is sure to change over time and will vary from place to place. Local data on reported incidence and estimates of under-reporting should be used to update this value. Similarly, population immunty will increase as the pandemic progresses. Sero-survey data can inform the proportion of the universiy community that is immune upon campus opening.

In conclusion, we present a model of SARS-CoV-2 transmission and control to assist universities in planning potential impacts and resource needs. Our model is conservative in that we assume a high reproductive number that is not reduced through non-pharmaceutical interventions. Despite this, we find that community-introduction of SARS-CoV-2 infection onto campus can be controlled with effective testing, isolation, contract tracing and quarantine, consistent with observations that this strategy has been successful in the general population where implemented properly (e.g. South Korea).(18) The results of this model simulation approach have been influential in Emory University’s decision to open in Fall 2020. The University will implement a comprehensive testing strategy and will shorten the semester with an early start, with no breaks in order to end by Thanksgiving, amongst a number of other strategies to suppress transmission.(19)

## Data Availability

Data and all underlying code available at https://github.com/lopmanlab/covid_campus_model

## Appendix I. Model equations

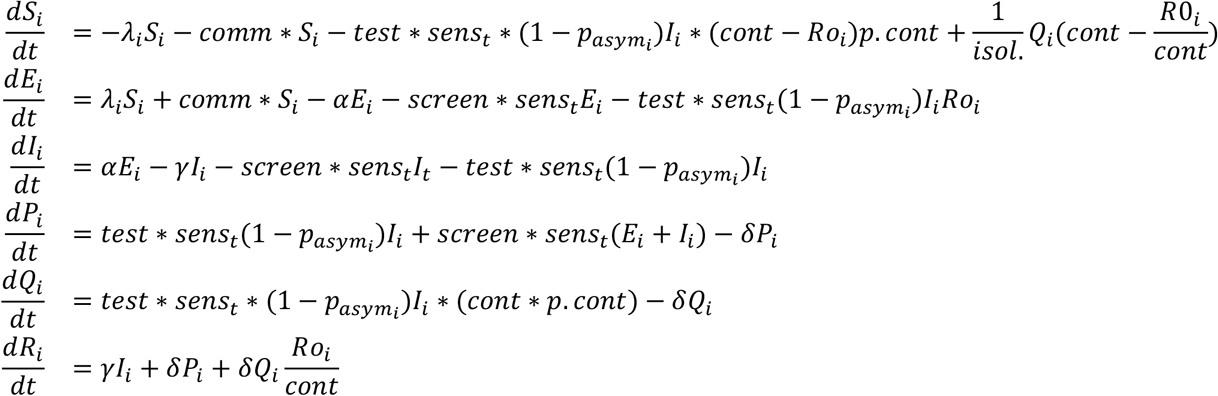

Where i &= on (students living on campus); off (students living off campus); saf (staff and faculty), and

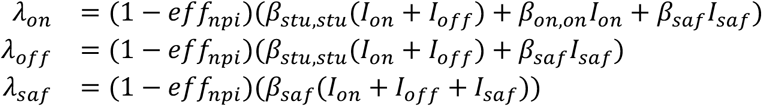

## Appendix II. Estimated active and cumulative cases under intervention scenarios with 25th and 75th centile range

## Appendix III. Partial rank correlation coefficient of key model inputs

## Notes

### Competing Interest Statement

The authors have declared no competing interest.

### Funding Statement

This work was supported by NIH/NIGMS R01GM12428003S1

